# A Joint Compartmental Model for The Co-infection of SARS-CoV-2 and Influenza

**DOI:** 10.1101/2022.08.26.22279281

**Authors:** Reyhaneh Zafarnejad, Paul M. Griffin, Mario Ventresca

**Affiliations:** Department of Industrial and Manufacturing Engineering, Penn State University, University Park PA 16803, USA; School of Industrial Engineering, Purdue University, West Lafayette, IN 47906 USA

**Keywords:** SARS-CoV-2, Influenza, Co-infection, Compartmental Modeling, Vaccination, Quarantining

## Abstract

Co-infection of COVID-19 and other respiratory pathogens, including influenza virus family, has been of importance since the beginning of the recent pandemic. As the upcoming flu season arrives in countries with ongoing COVID-19 epidemic, the need for preventive policy actions becomes more critical. We present a joint compartmental SEIRS-SIRS model for the co-circulation of SARS-CoV-2 and influenza and discuss the characteristics of the model, such as the basic reproduction number (R_0_) and cases of death and recovery. We implemented the model using 2020 to early 2021 data derived from global healthcare organizations and studied the impact of interventions and policy actions such as vaccination, quarantine, and public education. The VENSIM simulation of the model resulted in R_0_ = 7.5, which is higher than what was reported for the COVID-19 pandemic. Vaccination against COVID-19 dramatically slowed its spread and the co-infection of both diseases significantly, while other types of interventions had a limited impact on the co-dynamics of the diseases given our assumptions. These findings can help provide guidance as to which preventive policies would be most effective at the time of concurrent epidemics, and contributes to the literature as a novel model to simulate and analyze the co-circulation of respiratory pathogens in a compartmental setting that can further be used to study the co-infection of COVID-19 or similar respiratory infections with other diseases.

## 1. Introduction

The recent pandemic pathogen, Severe Acute Respiratory Syndrome Coronavirus-2 (SARS-CoV-2), which started in December 2019, has caused a wide range of illness varying from mild symptoms to complicated and severe respiratory response, and in 3% of cases even death^1–3^. Although at the time of writing this paper available data was limited, recent case reports of concurrent infection of influenza virus in adults and children with SARS-CoV-2 infection have suggested that co-infection may heavily influence morbidity and mortality^4^. Previous literature has shown that co-infections were frequent in patient populations and is of importance for both flu season and deadly variants of SARS-CoV-2 ^5–8^. The Centers for Disease Control and Prevention (CDC) has reported that influenza and COVID-19 have overlapping signs and symptoms, and co-infection has been documented in both case reports and case series^9^.

The reported impact of existing infection with SARS-CoV-2 and co-infection with other pathogens varies, from negative^10^ to not significant^11^ to positive^12,13^. For example, Kim et al. ^11^ reports more than 20% of 116 SARS-CoV-2 positive individuals tested over a 20 day period contained one or more additional respiratory pathogens, most often rhinovirus/enterovirus and different types of influenza virus family, although the prevalence of co-infection among COVID-19 positive and negative population was not statistically significant. In another study, Yue et al.^12^ showed that the prevalence of co-infection with influenza among a group of COVID-19 positive patients in Wuhan, China was more than 50%, while the prevalence of infection with influenza virus pre-pandemic was less than 1%. In a recent study, Bai et. al^13^ found that influenza A virus (IAV) pre-infection significantly promoted the infectivity of SARS-CoV-2 in a broad range of cell types through an experimental co-infection with IAV and SARS-CoV-2 virus.

In terms of preventive policy actions during the recent pandemic, policy makers across the globe designed different strategies to try to control the pandemic ^14^. In the early stages of the pandemic, while pharmaceutical interventions such as vaccination and medical treatment were not accessible, non-pharmaceutical interventions were widely implemented^15^. These interventions, such as mask mandates, social distancing, quarantining, surveillance testing, and contact tracing were substantially effective in slowing down the progression of COVID-19^16–18^ and some of them are still in place to date. These non-pharmaceutical interventions were found to reduce the burden of flu as well^19^, intuitively due the similarities in respiratory propagation of both viruses and how the parallel spread of diseases were slowed down by general hygiene enforcement and social contact reduction. Vaccination programs for COVID-19, becoming accessible worldwide midway through the pandemic, played a significant role in reducing the spread of COVID-19, and associated hospitalization and mortality^20^. With the emergence of new variants of COVID-19, vaccination proved to be as impactful although booster doses were required and remain ongoing globally^21^. However, with the rise of new variants, achieving herd immunity still remains out of reach ^22^. Nevertheless, flu shots are promoted by public health officials and are found to be effective in controlling simultaneous outbreak of influenza and COVID-19^23^. However, it is unclear how impactful pharmaceutical interventions are in reducing co-infection cases of COVID-19 and influenza.

Understanding the co-existence and co-infection of two or more diseases at the same time has been an important and controversial topic in the field of epidemiology. To name a few, in 2016, Naji and Hussein^24^ proposed a compartmental model describing the dynamics of the spread of two different types of pathogens based on two underlying models of disease spread, an SIS-type disease and an SIRS-type disease. In another study, Tilahun et. al^25^ studied the co-dynamics of Pneumonia as an airborne disease and Typhoid fever as a vector-based disease using a joint SIRS-SIRS simulation for cost-effective disease control purposes. More recently, Rehman et. al^26^ developed a mathematical transmission model for the co-infection of dengue fever and COVID-19, and described the co-dynamics of the propagation using qualitative and numerical analysis. However, there is limited literature on modeling the co-existence and co-circulation of SARS-CoV-2 and influenza viruses as two airborne diseases.

In this paper, we describe a model of disease progression consisting of two joint compartmental models for COVID-19 and influenza. We further simulate the behavior and transmission dynamics of diseases based on the most recent data including US COVID-19 updates and CDC and WHO guidelines. Moreover, we discuss how our results compare and contrast with observed data and published literature since the COVID-19 pandemic started. Finally, we study the impact of vaccination against COVID-19, and non-pharmaceutical interventions such as education and social distancing on the behavior of COVID-19 – influenza co-circulation. The results of this study could provide useful information for researchers and policy makers as to which policy action could mitigate the negative impacts of concurrent epidemics on mortality rate and total cases of infection. With the possibility of emergence of other respiratory epidemics and pandemics in the future^27^, mathematical and predictive models of co-infection with multiple diseases could further be used in cost-effectiveness and cost-utility analyses and help develop adaptive and effective policy actions.

## 2. Materials and Methods

We proposed a SEIRS-SIRS compartmental model to study the co-existence of COVID-19 and influenza and further implemented the Next Generation Method (NGM) to find the basic reproduction rate (R_0_). We also performed sensitivity analysis and solved the model for various parameter values in order to understand the effect of interventions and policy actions on the spread of diseases ^28^. We created a simulation model for validation and additional investigations.

### 2-1. Model Parameters and Relationships

To capture the co-dynamics of COVID-19 and influenza, we developed a joint SEIRS-SIRS compartmental model (Figure 1). This model considers a population (N) that is divided into nine compartments, susceptible (S), COVID-19 exposed (E_c_), COVID-19 infectious (I_c_), influenza infectious (I_f_), COVID-19 and influenza co-infectious (I_fc_), COVID-19 and influenza co-exposed (E_fc_), COVID-19 recovered (Rc), influenza recovered (R_f_) and COVID-19–influenza co-infectious recovered (R_fc_). We assumed a closed environment with birth rate and natural death rates both equal to 0, while the number of susceptible population increases by those individuals that lose their temporary immunity^29^ from the recovered compartment of COVID-19 (R_c_), influenza (R_f_) and COVID-19–influenza co-infected compartment (R_fc_) with rates of d_1_, d_2_ and d_3_, respectively.

**Figure 1.**
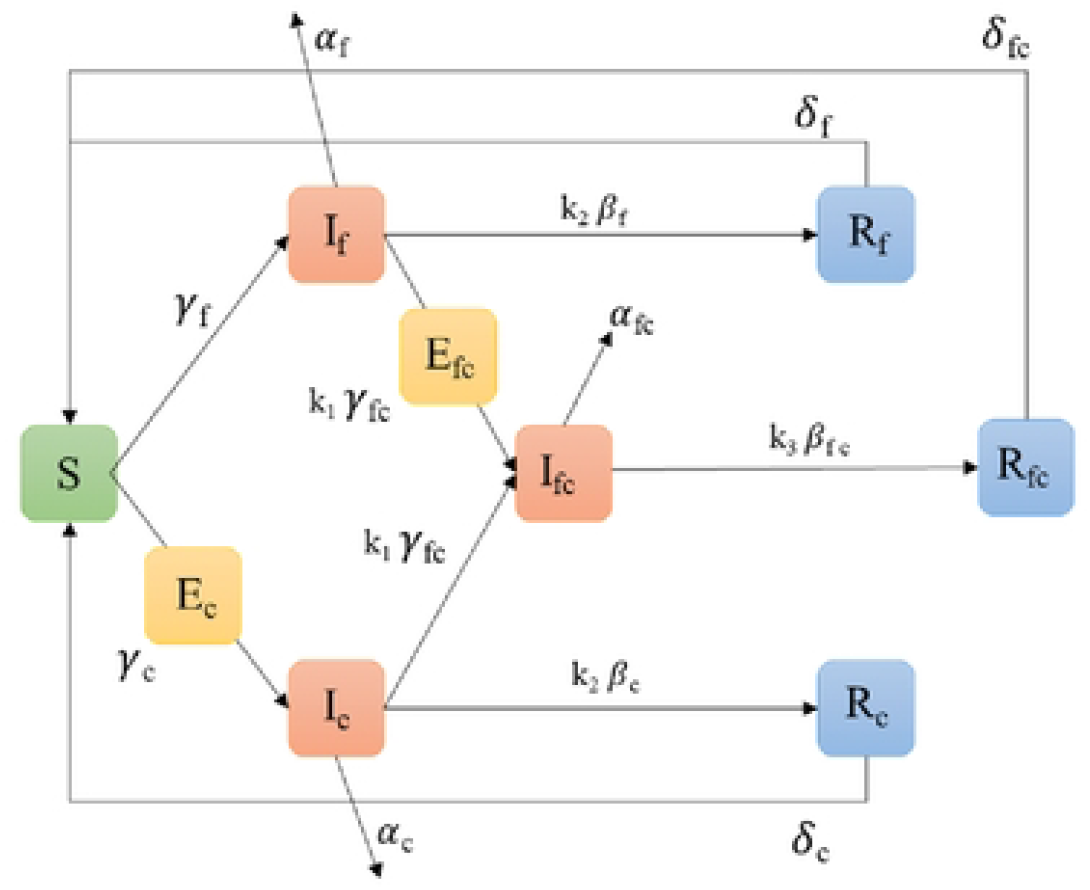
Schematic flow of the model, parameters and compartments are outlined in section 4.1 and dynamics are described in section 4.2. Different colors are utilized to indicated similar components. Figure 1 illustrates the model described in Section 4.1. The corresponding dynamics are provided in Equations 1 - 10.

Susceptible individuals become infected either with COVID-19 at the rate *γ*_*c*_ and join the COVID-19 exposed compartment (E_c_), or with influenza at infection rate *γ*_*f*_ and joining influenza infectious compartment (I_f_). Patients that are exposed to COVID-19 will become infectious after the latent period (L_c_). Since the latent period for influenza is relatively low (roughly from 0.64^30^ days to 1.6 days^31^), we ignored the latency period for influenza. Similarly, patients already infected with influenza might get infected with COVID-19 with the co-infection rate *γ*_*fc*_ and become COVID-19 exposed, and after the latent period become co-infected.

The infectious compartment of COVID-19 can receive treatment or recover naturally at the rate β_c_ and move to COVID-19 recovered compartment (R_c_) or die with a death rate of α_c_. Similarly, the infected compartment of influenza can receive treatment or recover with a rate of β_f_ and join the influenza recovered compartment (R_f_) or die at a rate of α_f_. Moreover, the COVID-19-influenza co-infected compartment transitions from the co-infected compartment to the recovered compartment with a rate of β_fc_ and obtain temporary immunity and therefore join the co-infected recovered compartment (R_fc_). We assumed that all recovered compartments tend to become susceptible once again, after a specific amount of time (*δ*_f_, *δ*_c_ and *δ*_fc_ for flu recovered, COVID-19 recovered and co-infection recovered, respectively).

An important assumption we made is considering adjustment parameters, k_1_, k_2_ and k_3_ as additional coefficients in the model. Since this model is essentially the combination of two disjoint SEIRS and SIRS compartmental models, overlaps are inevitable. Patients might technically belong to more than one compartment in reality, but these models are unable to easily capture this complexity. For the model to produce valid results, we assumed that only a proportion of COVID-19 or influenza infected individuals are at risk for co-infected with the other pathogen, and therefore assigned adjustment probabilities^25,26^. These additional parameters control the flow between compartments and adjust the daily per capita co-infection rate and recovery rates so that the resulting trends mimic real world observations. Adjustment parameters received values between 0 and 1 with the sum of 1.

### 5-1. Mathematical Model and adjustments

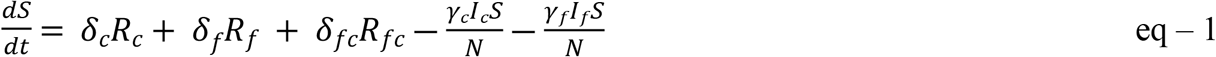

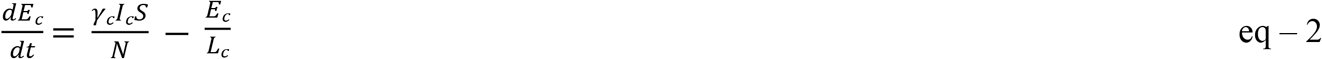

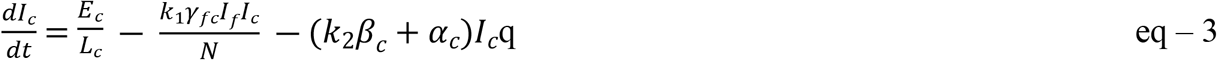

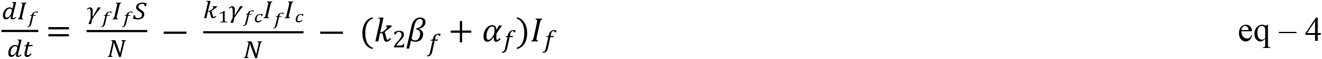

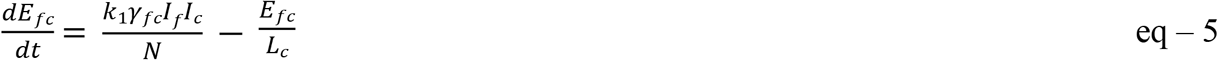

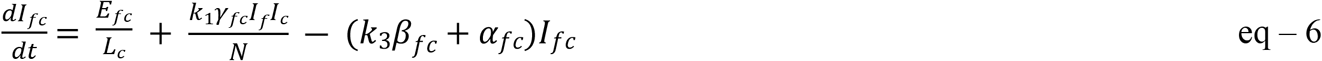

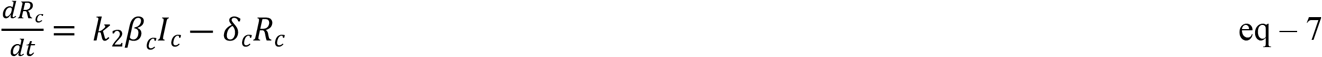

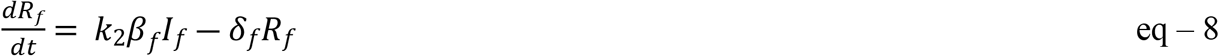

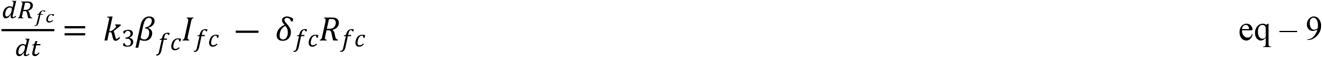

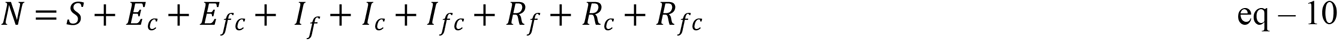

### 2-2. Basic Reproduction Number (R_0_)

The Basic Reproduction Number (R_0_) is used to measure the transmission potential of a disease and is equal to the average number of secondary infections produced by a typical case of an infection in a population where everyone is susceptible^32,33^. The next generation method is used to calculate the R_0_ associated with the model of co-infection of SARS-CoV-2 and influenza virus^34^. This system has five infected states: E_c,_ E_fc_, I_c_, I_f_ and I_fc_; and four uninfected states: S, R_c,_ R_f_ and R_fc_. Although there are nine states in the model, it is eight-dimensional as the total population size is constant. At the infection-free steady state, E_c_ = E_fc_ = I_c_ = I_f_ = I_fc_ = R_c_= R_f_ = R_fc_ = 0, hence S = N. Therefore, for small (E_c,_ E_fc_, I_c_, I_f_, I_fc_) we have the following non-linear system

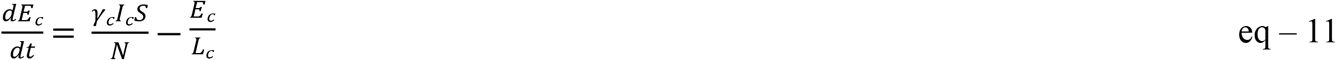

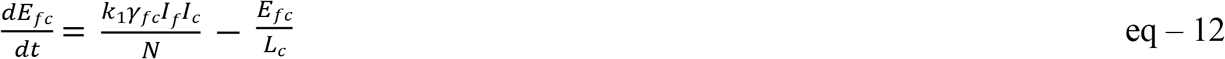

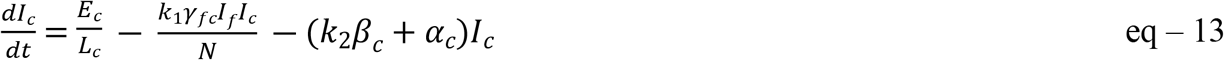

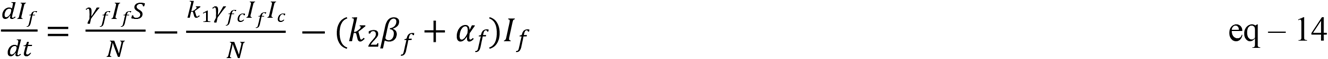

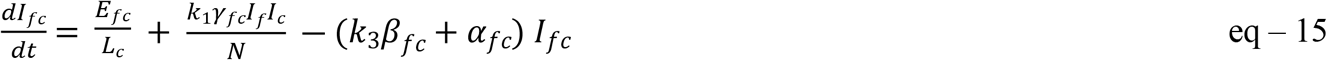

The transition (F) and transmission (V) matrices are as follows:

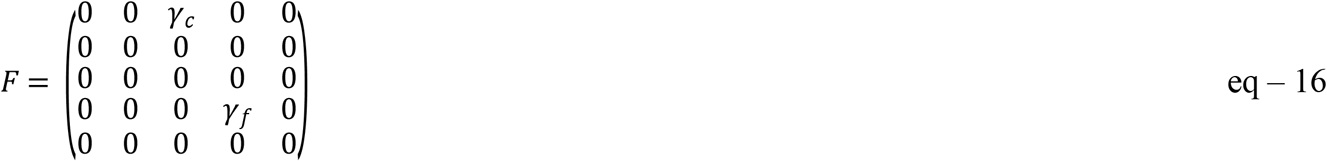

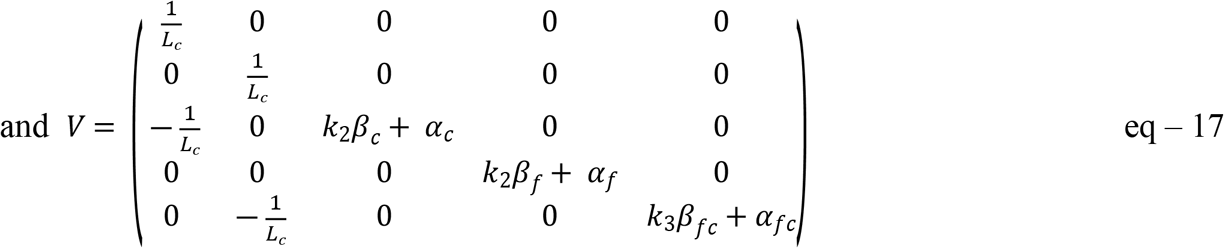

The eigenvalues of *FV*^−1^ are obtained as:

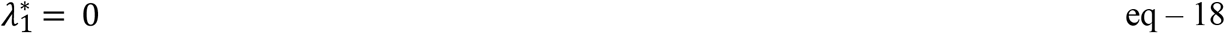

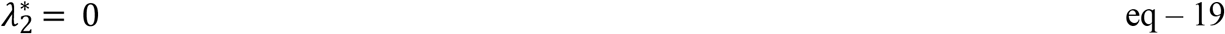

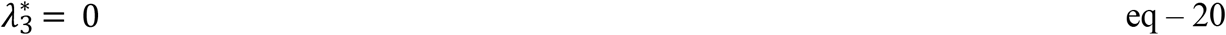

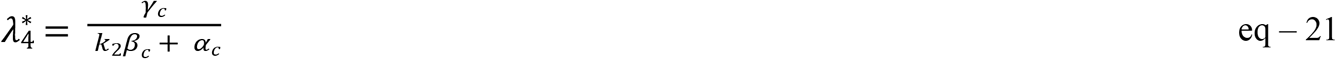

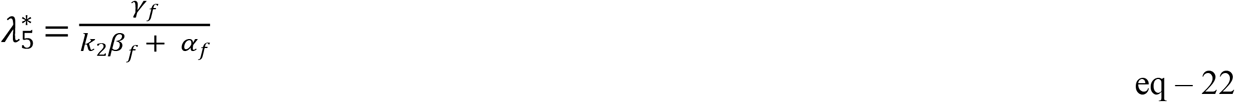

The basic reproduction number is computed as the spectral radius of *FV*^−1^

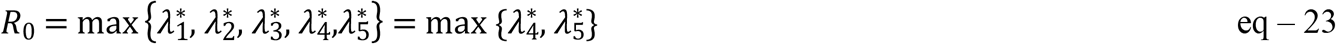

which after simplification is independent from the co-infection rate (*γ*_*fc*_) and co-infection recovery rate (*β*_*fc*_), and depending on the evaluating parameters, the dominant R_0_ is merely determined by either COVID-19 or influenza branch. We discuss the implications of R_0_ in Section 2-2

### 2-3. Simulation

In order to study the co-spread of COVID-19 and influenza based on the proposed model, we developed a system dynamics simulation model in VENSIM software V 8.0.9^35^. The full model is provided in Appendix 1 and more details will be provided on request. Table 1 summarizes the parameter values and available references implemented in the simulation. We modeled the co-infection of SARS-CoV-2 for the state of Indiana with the population of 6,732,000 according to 2019 US census^36^. The parameter values are derived form 2020 available COVID-19 dashboards and databases. We made necessary assumptions in cases where convenient or proper data was not accessible, the most important of which is the estimation of co-infection rate. As discussed previously in the Introduction, it is still unclear whether infection with COVID-19 or other respiratory pathogens affects the co-infection rate. By roughly estimating the average of values reported in the literature, we assumed that the daily per person co-infection rate is 20% more than the maximum of infection rate with either COVID-19 or influenza. The rest of the parameters, references and assumptions are provided in Table 1.

**Table 1.**
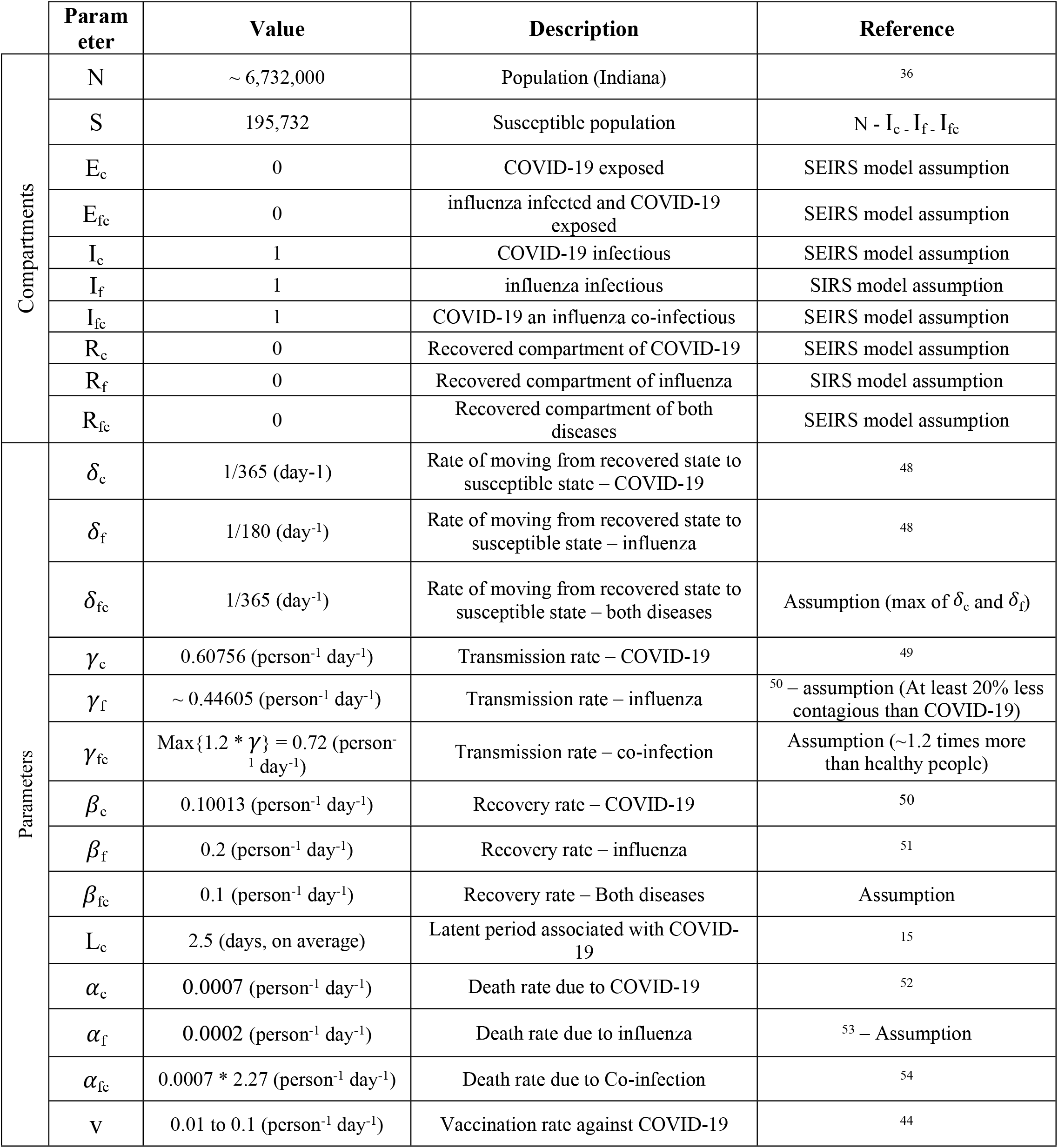
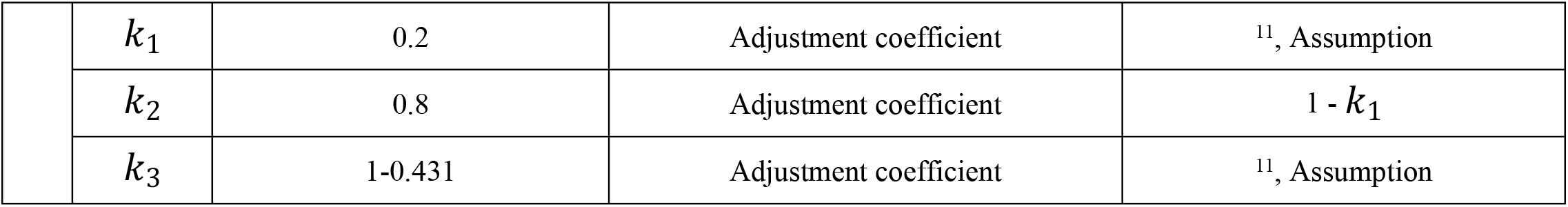
Model parameters summary

|  | Parameter | Value | Description | Reference |
| --- | --- | --- | --- | --- |
| Compartments | N | ~ 6,732,000 | Population (Indiana) | 36 |
| | S | 195,732 | Susceptible population | $N - I_c - I_f - I_{fc}$ |
| | $E_c$ | 0 | COVID-19 exposed | SEIRS model assumption |
| | $E_{fc}$ | 0 | influenza infected and COVID-19 exposed | SEIRS model assumption |
| | $I_c$ | 1 | COVID-19 infectious | SEIRS model assumption |
| | $I_f$ | 1 | influenza infectious | SIRS model assumption |
| | $I_{fc}$ | 1 | COVID-19 an influenza co-infectious | SEIRS model assumption |
| | $R_c$ | 0 | Recovered compartment of COVID-19 | SEIRS model assumption |
| | $R_f$ | 0 | Recovered compartment of influenza | SIRS model assumption |
| | $R_{fc}$ | 0 | Recovered compartment of both diseases | SEIRS model assumption |
| Parameters | $\delta_c$ | 1/365 (day<sup>-1</sup>) | Rate of moving from recovered state to susceptible state – COVID-19 | 48 |
| | $\delta_f$ | 1/180 (day<sup>-1</sup>) | Rate of moving from recovered state to susceptible state – influenza | 48 |
| | $\delta_{fc}$ | 1/365 (day<sup>-1</sup>) | Rate of moving from recovered state to susceptible state – both diseases | Assumption (max of $\delta_c$ and $\delta_f$) |
| | $\gamma_c$ | 0.60756 (person<sup>-1</sup> day<sup>-1</sup>) | Transmission rate – COVID-19 | 49 |
| | $\gamma_f$ | ~ 0.44605 (person<sup>-1</sup> day<sup>-1</sup>) | Transmission rate – influenza | <sup>50</sup> – assumption (At least 20% less contagious than COVID-19) |
| | $\gamma_{fc}$ | $\text{Max}\{1.2 * \gamma\} = 0.72$ (person<sup>-1</sup> day<sup>-1</sup>) | Transmission rate – co-infection | Assumption (~1.2 times more than healthy people) |
| | $\beta_c$ | 0.10013 (person<sup>-1</sup> day<sup>-1</sup>) | Recovery rate – COVID-19 | 50 |
| | $\beta_f$ | 0.2 (person<sup>-1</sup> day<sup>-1</sup>) | Recovery rate – influenza | 51 |
| | $\beta_{fc}$ | 0.1 (person<sup>-1</sup> day<sup>-1</sup>) | Recovery rate – Both diseases | Assumption |
| | $L_c$ | 2.5 (days, on average) | Latent period associated with COVID-19 | 15 |
| | $\alpha_c$ | 0.0007 (person<sup>-1</sup> day<sup>-1</sup>) | Death rate due to COVID-19 | 52 |
| | $\alpha_f$ | 0.0002 (person<sup>-1</sup> day<sup>-1</sup>) | Death rate due to influenza | <sup>53</sup> – Assumption |
| | $\alpha_{fc}$ | $0.0007 * 2.27$ (person<sup>-1</sup> day<sup>-1</sup>) | Death rate due to Co-infection | 54 |
|  | v | 0.01 to 0.1 (person<sup>-1</sup> day<sup>-1</sup>) | Vaccination rate against COVID-19 | 44 |

|  |  |  |  |
| --- | --- | --- | --- |
| $k_1$ | 0.2 | Adjustment coefficient | <sup>11</sup>, Assumption |
| $k_2$ | 0.8 | Adjustment coefficient | $1 - k_1$ |
| $k_3$ | 1-0.431 | Adjustment coefficient | <sup>11</sup>, Assumption |

### 2-4. Interventions

The simulation was analyzed for three modified versions of the model to capture the impact of different intervention settings. The primary model (i.e. the baseline model) assumes COVID-19 and influenza propagation began simultaneously at T=0. The second model includes vaccination against COVID-19 as a pharmaceutical intervention, and the third model considers the impact of non-pharmaceutical interventions, such as quarantining, public education and social distancing. Although the flu occurs mostly as a seasonal disease, for the sake of simplicity we assumed simultaneous propagation of both COVID-19 and influenza. We simulated the co-circulation of COVID-19 and influenza for 365 days, for an initial population of S_0_ = 6,732,000. The initial value for all other compartments was assumed to be 0 at time T=0, except for the infected compartment which initially contains 1 patient (local patient zero).

In terms of the mathematical structure of the model, the impact of vaccination and quarantining/public education is described as follows. Vaccination against COVID-19 increases the rate of exit from the susceptible subgroup:

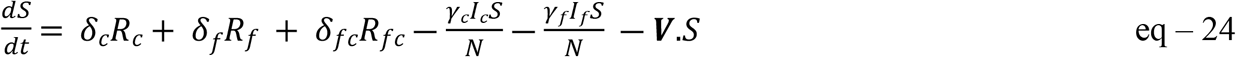

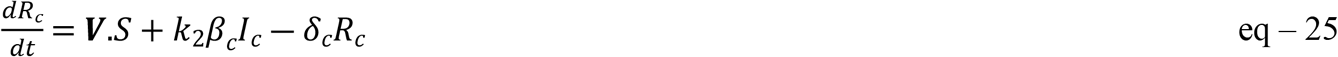

where V indicates the vaccination rate (person^-1^ day^-1^)

Quarantining/public education on the other hand affects the rate of infection with COVID-19, influenza as well as the rate of co-infection, through decreasing the contact rate between individuals:

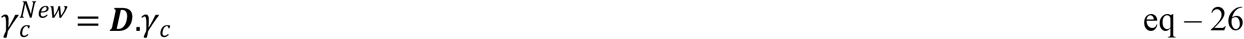

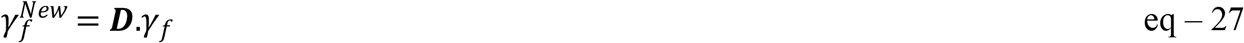

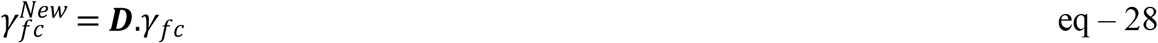

where D indicates the decrease in contact rate. The rest of the mathematical model remains unchanged.

## 3. Results

### 3-1. Baseline Model Simulation Results

Figure 2 shows an overview of all the compartments present in the model over the course of the simulation, as well as a detailed comparison between compartments. The parameter values were derived from 2020 and early 2021 national and global databases. As demonstrated in Figure 2 and the corresponding subfigures, in the base model with no interventions, the peak for infection of COVID-19 was significantly higher than for influenza and co-infection of both. The three peaks almost occurred at the same period, with a lower peak for the influenza in comparison to COVID-19 and a dramatically lower peak for co-infection in comparison to the other types of infection. Despite similarities between SARS-CoV-2 and influenza virus family, the transmission rate associated with COVID-19 was at least 20 percent (to up to 3 times) higher than influenza. This is why we see a clearly smaller peak of infection associated with influenza.

**Figure 2.**
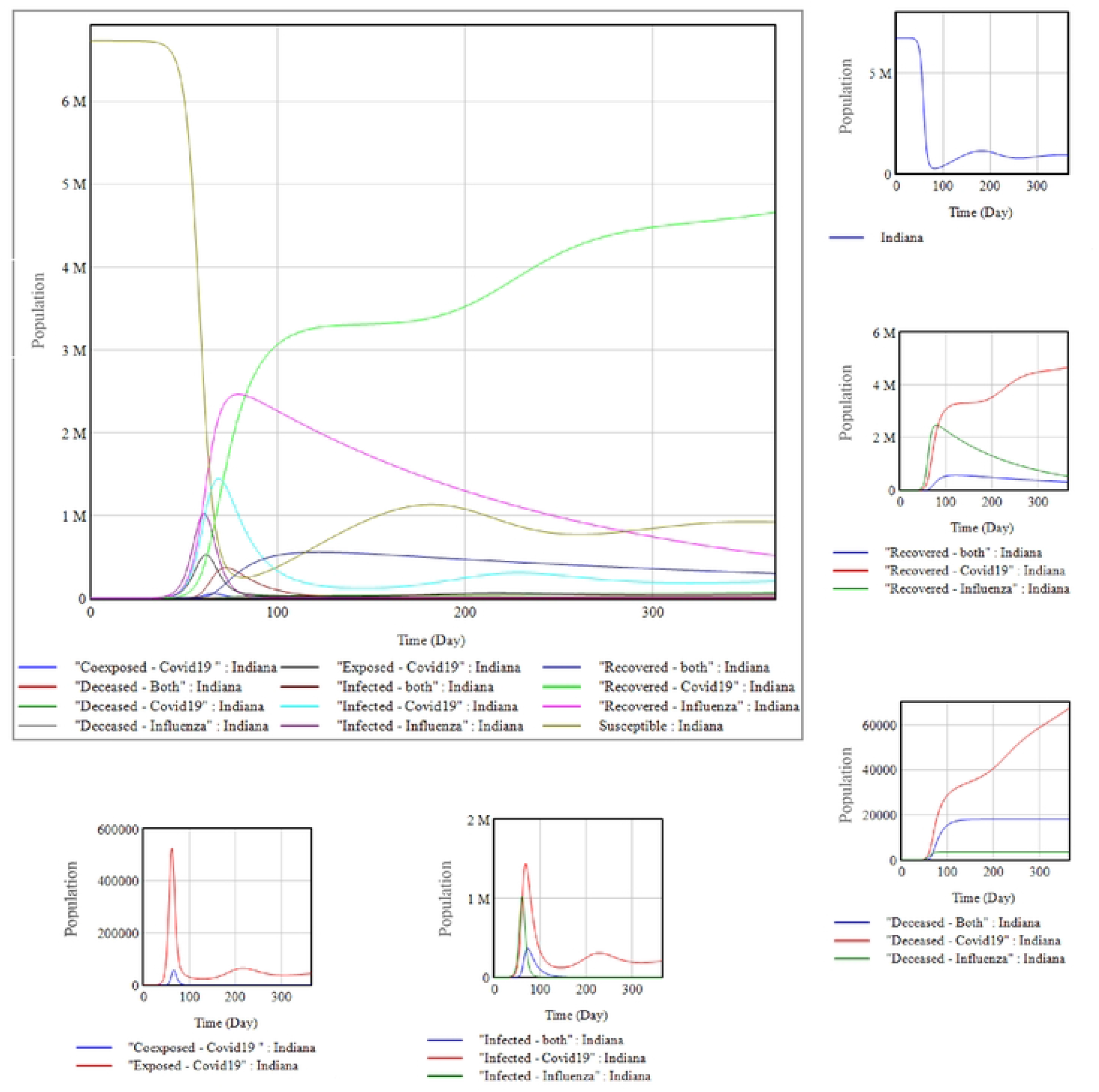
An overview of the compartments in the model. The smaller figures illustrate a closer comparison between similar compartments, for a 365-day simulation of the Co-infection of COVID-19 and Influenza.

A second and smaller peak in the COVID-19 infected population occurred after around 150 days of the first peak, which is in accordance with previously published data for the US from February 20, 2020 to December 21, 2020^37^. No such behavior was found in association with influenza, which also agrees with the expected behavior of annual influenza epidemy in Indiana population^38^ prior to any COVID-19 related interventions.

On the other hand, the recovered compartment of COVID-19, influenza, and co-infection of both behave differently over time. Three existing recovered compartments (recovered from COVID-19, influenza, or the co-infection of both), experienced a peak at around the same time (COVID-19 is behind due to the incubation period), similar to the infected compartment with the COVID-19 recovered population with a significantly higher peak (due to more cases of infection). We can see that based on this model for a 365-day simulation, after around 9 months almost exclusively COVID-19 remains active and contagious, particularly after the second wave.

The susceptible and deceased compartments also showed interesting underlying behaviors over the course of the simulation. Over the first year of pandemic, over 96% of the susceptible population suffered from at least one of the infections which is relatively high. The smaller rise in the susceptible population occurred exactly before the second wave of the COVID-19 pandemic (T = 229 days). This indicates that before the second wave, the number of individuals in the susceptible compartment increased due to recruitment of susceptible individuals by recovery. Consequently, the second wave appeared, and an additional number of infected individuals led to another less steep fall in the number of susceptible individuals. The same behavior was observed for the number of deceased populations, more COVID-19 infected individuals in the second wave led to more deaths due to COVID-19, while the number of deaths caused by influenza remained the same (∼3481), with a very slight increase for individuals infected with both diseases (∼100 additional deaths).

The simulated death rates and observed trends also mimic recent observations both in the US in general and Indiana^37^, as the first and second wave in the total COVID-19 deaths during 2020 indicates how well this model compares with the published literature.

### 3-2. Calculation of R_0_

Given the information and parameter values provided in Table 1, the basic reproduction number (R_0_) can be estimated for the model based on Eq. 23. In this case, we have

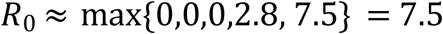

which in comparison to the median R_0_ associated with COVID-19 in the US (5.8 - 95% CI 3.8– 8.9)^39,40^ in 2020 and early 2021, as well as R_0_ associated with seasonal flu (1.2 – 1.3)^41^ indicates higher contagiousness of COVID-19 and influenzas co-circulation.

### 3-3. Interventions and Policy Actions

To model vaccination against COVID-19, we assumed a transition between the susceptible compartment and the COVID-19 recovered compartment. Individuals in the susceptible group can either get infected by COVID-19 or influenza, or directly go to COVID-19 recovered compartment based on a vaccination rate. Therefore, by running sensitivity analysis on the model with various vaccination rates we estimated the impact of vaccination on co-circulation of infections. On the other hand, quarantine and education of people both were assumed to reduce the number of contacts, mostly among the susceptible population, resulting in a reduction in the rates of infection (*γ*). In other words, we studied two types of interventions: those affecting the number of contacts per person per day, and those affecting the transition between stages by adding new transition states.

#### 2-3-1. Vaccination against COVID-19

By April 2021, the US has administered more than 3 million vaccine shots per day, or around 0.01 shots per person daily across the country^33^. To study the impact of vaccination against COVID-19, we defined a varying range for vaccination rate for the state of Indiana, ranging from 0 to 0.005 (person per day) and further ran sensitivity analysis on the impact of vaccination rate on the model. Figure 3 demonstrates how sensitive the size of each compartment is to the rate of vaccination. As the rate of vaccination increased, a visible decrease in the number of infected individuals could be identified; daily flu infection and COVID19 – flu co-infection cases decline for at least 85% as the daily per capita vaccination rate increases by 0.0005 person.day^-1^. This in fact is in accordance with the minute level of reported flu cases in 2020 and early 2021^42^. Similarly, yet more slowly for the COVID-19 daily case, with a 0.001 increase in daily per capita vaccination rate there was a 45% reduction in the total number COVID-19 infected individuals. The second wave of COVID-19 was also mitigated as the vaccination rate increased. The total number of deaths also decreased with the higher prevalence of vaccination. The impact of vaccination was more pronounced on the influenza and co-infection compartments, which can be justified due to lower incubation rate associated with influenza in comparison to COVID-19.

**Figure 3.**
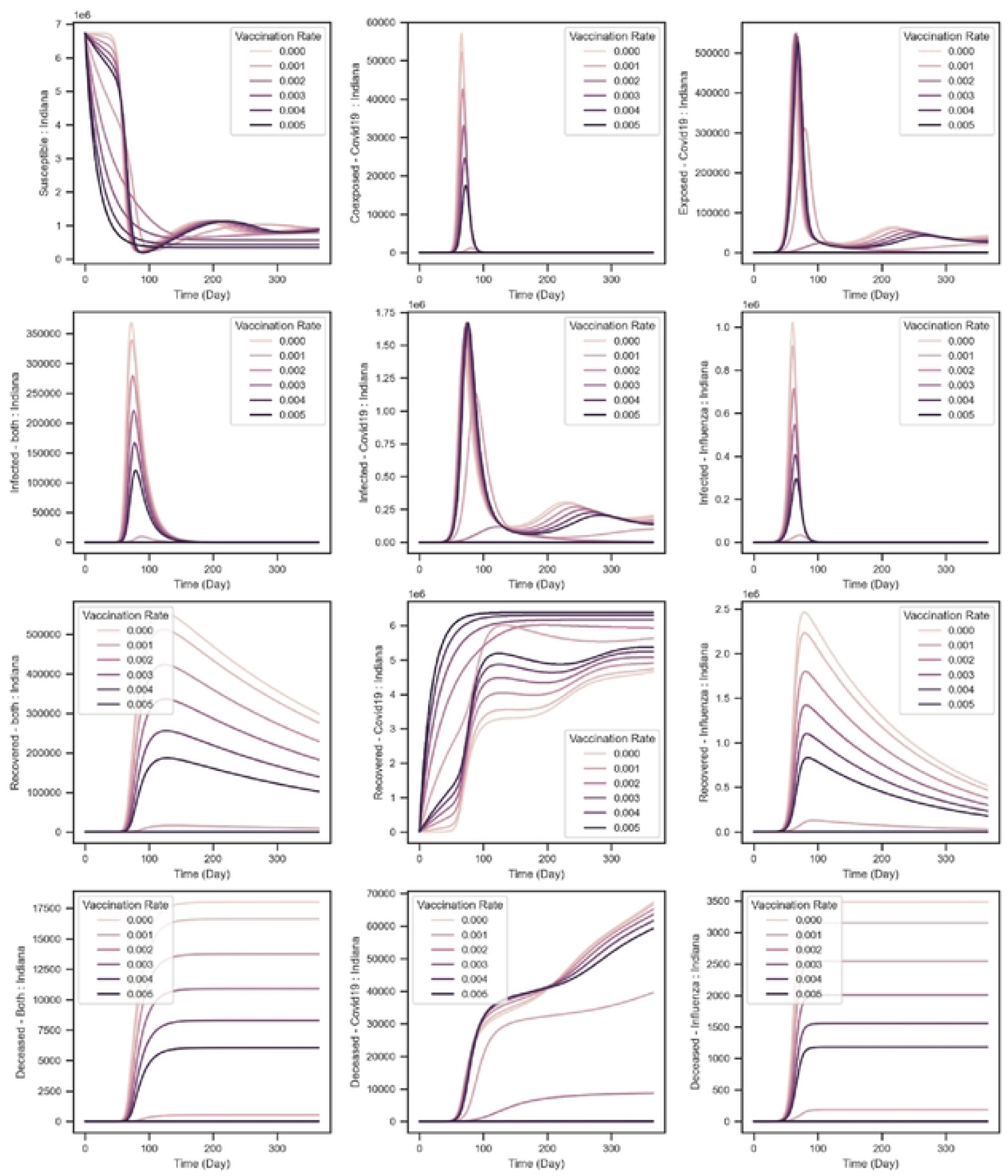
The effect of COVID-19 vaccine on the simulation results – lighter curves indicate smaller rate of vaccination.

An important point to consider here is the fact that, in this setting, higher vaccination rates corresponded to faster transitions between the susceptible compartment and the COVID-19 recovered compartment. As mentioned previously, from the modeling perspective, vaccination plays the role of a shortcut from susceptible compartment to recovered compartment. That is why there is a fall in the number of susceptible individuals as the rate of vaccination against COVID-19 increases. This also affects the total number of COVID-19 recovered individuals, which is not entirely due to infection.

#### 2-3-2. Quarantine and education

In order to study the impact of non-pharmaceutical interventions such as quarantine and education^43^, we assumed that the contact rate per person can decrease by up to 80% by quarantining non-infected individuals, social distancing and educating the public. This assumption is made to enable the model to capture a wide range of contact rates, without considering the feasibility of providing so in practice. In this case, the simulation structure remains the same as the original simulation except for the values of *γ*_*f*_, *γ*_*c*_ and *γ*_*fc*_ will be reduced by up to 20% of the original values, in increments of 10 percent. As shown in Figure 4, as the contact rate declined from 1 (100%) to 0.2 (20%), the total number of deceased individuals dropped dramatically in all the deceased compartments. Similarly, the peak for influenza infection and COVID-19 – influenza infection dampened as the number of contacts decreased. For example, a 50% reduction in the contact rate resulted in more than a 90% reduction in the daily cases of influenza and co-infection and led to a delay of around 75 days in the peak of COVID-19 propagation. However, in the case of individuals with COVID-19 only, the primary effect of the reduction in contact rate was a delayed peak. As the contact rate decreased (from 70% of the maximum effective contact rate to 20%), we observed more than 60% reduction in the peak of COVID-19 cases as well.

**Figure 4.**
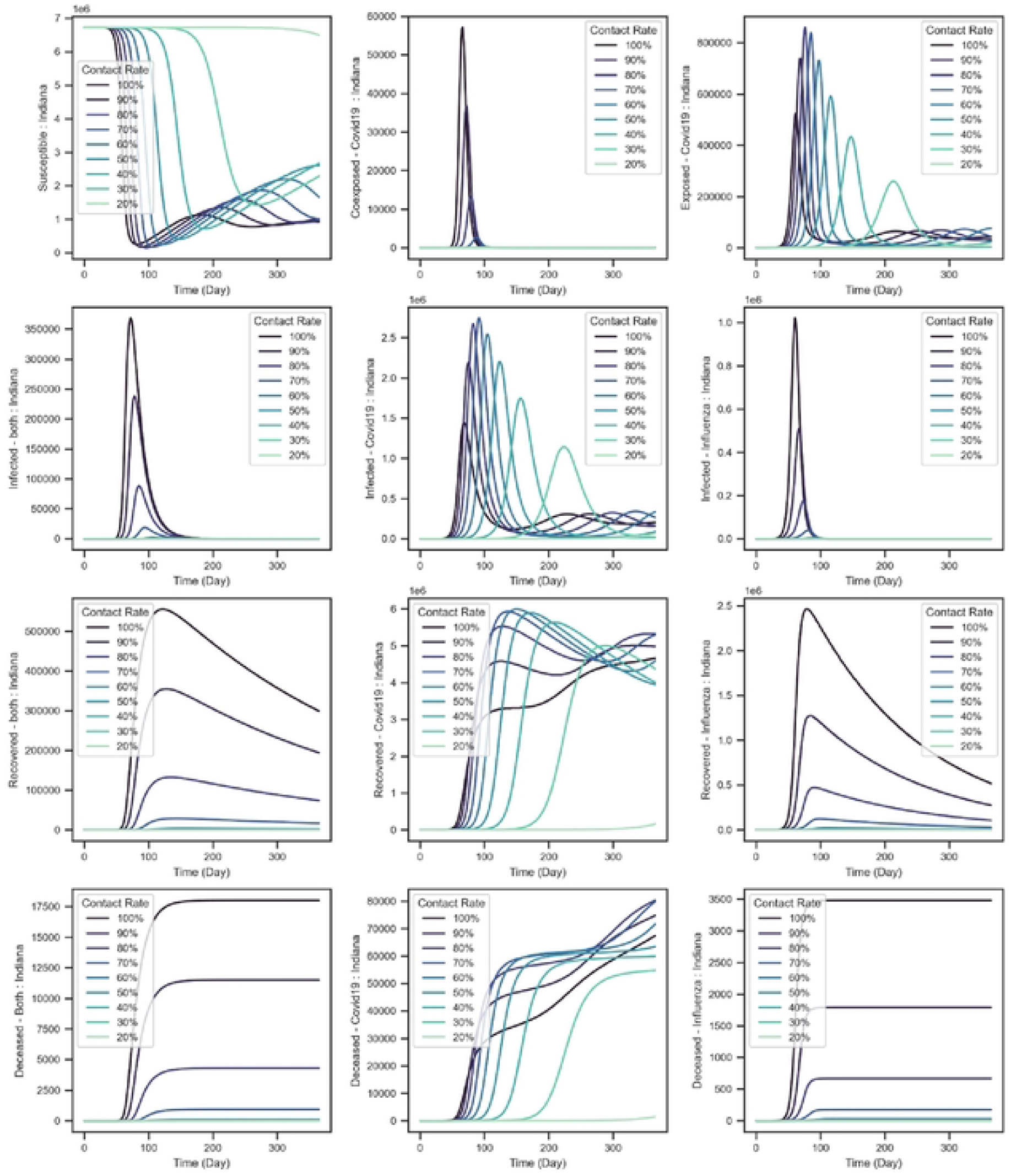
The effect of quarantine and education on the simulation results – lighter curves indicate higher decrease in the contact rates (1: no decrease in contact rates, 0.2: 20% of the maximum effective contact rate, etc.)

Like what was observed through vaccination, changes in the contact rate induced similar behavior in the model, such as the short-term increase (for a window of about 100 days) in the peak of COVID-19 infection. Similar behavior was found in other compartments of COVID-19 infection branch as well, and in all cases the peak occurred later in time. We can explain this behavior based on both the model structure and assumptions. Here we noticed a steady reduction in influenza infection cases and the number of cases in other influenza related compartments, which indicates the sensitivity of influenza to the contact rate, unlike COVID-19 with a short term increase and further decrease behavior. Since fewer patients moved from the susceptible compartment to the influenza infected compartment (up to 60% less in comparison to COVID-19 cases at the peak), more individuals would remain susceptible and further move to the COVID-19 exposed and infected compartments. This is a weakness of compartment models per se, which prevents individuals from belonging to multiple compartments at the same time. Therefore, we cannot rely on these results to estimate how well education and quarantining can reduce the speed of infection. However, we can claim that reducing the contact rate was associated with delay in when the peak in infection curve occurred.

## 4. Discussion

In this paper, the co-circulation of SARS-CoV-2 and influenza was studied using a joint SEIRS-SIRS compartmental model, including the impact of various interventions and policy actions. In addition, the basic reproduction number associated with the co-infection of COVID-19 and influenza was computed and found to be heavily influenced by the COVID-19 branch. The basic reproduction number computed based on the model (∼7.5) was also higher in comparison to R_0_ associated with COVID-19 or influenza separately, pointing to the importance of understanding and mitigating co-infection.

In terms of interventions, the effect of interventions such as vaccination against COVID-19 was found to be significant in controlling the spread of COVID-19 alongside seasonal flu, with more than a 65% reduction in the total number of deceased individuals per year based on a vaccination rate of 3 susceptible individuals out of 1000 per day. Other types of interventions that affect the rate of transmission were not as successful as vaccination in reducing the total number of infected and deceased individuals but did effectively delay the peak in infection. Both interventions resulted in significant decrease in the number of flu cases, which is what was observed in 2020^42^. This implies that preventive interventions against COVID-19, pharmaceutical or non-pharmaceutical, automatically reduced the effect of other circulating respiratory pathogens such as influenza virus family.

There are several limitations to our work. For the sake of simplicity, we made several assumptions such as negligible incubation period for influenza, adjustment coefficients, and vaccination only against COVID-19, and parameterized the model with data from the developing, and in some cases contradicting, literature on the ongoing COVID-19 pandemic. Moreover, we limited the simulation to a closed environment with natural rates of birth and death equal to zero, as well as a 365-day simulation period that could lead to missing patterns of disease propagation and multiple waves that might occur over a period longer than a year. The model also only accounts for simultaneous co-infection and not sequential infection for periods shorter than 6 months (the recovery period of influenza) which could lead to biased simulation results. Another limitation to this work is that the influenza virus family was considered to be a seasonal infection, usually occurring in the fall and winter^44^ with a year-round circulation (with an expected peak between December and February in the US). On the other hand, there is lack of agreement in the literature over whether infection with influenza can potentially block infection with COVID-19^13,45,46^, in this model we only focused on the possibility of higher co-infection rates among patients infected with other pathogens. Further research could increase the accuracy of the current model by relaxing some of those assumptions.

Despite these limitations, our model successfully mimicked the observed patterns of COVID-19 and flu infections throughout 2020 and early 2021, when the COVID-19 pandemic was still away from an endemic phase. For future work, considering a simple cost-and-effect analysis, this model can assist healthcare policy makers to design and establish more efficient and less costly interventions to control the co-spread of such diseases^36^. In the times of deadly pandemics such the recent COVID-19 pandemic, making cost-effective decisions regarding control policies can heavily determine how well the economy can tolerate the impacts associated with such chaotic situations.

## 5. Conclusion

The simulation analysis presented in this work could provide public health officials with modeling tools and information that will help them to issue proper preventive guidelines and policy actions for the upcoming flu season in the southern hemisphere, and particularly in countries with yet ongoing COVID-19 crisis. Moreover, this work contributes to the current literature by introducing a novel epidemic model for simulating the co-dynamics of respiratory infection with two or more infectious pathogens, and has applicability in other settings including co-infection of Sexually Transmitted Diseases (STDS), Human Immunodeficiency Virus (HIV), Bacterial Pneumonia, etc.

## Data Availability

The VENSIM simulation models used during the current study are available and can be found at https://github.com/Rey-Zafarnejad/A_Joint_Compartmental_Model_for_The_Coinfection_of_SARSCoV2_and_Influenza. Further details on the model and simulation can be provided upon request.

## Author contribution

RZ helped define the problem, helped develop the model, ran the simulations, and helped in the writing of the manuscript. MV helped define the problem, helped interpret the results, and helped in the writing of the manuscript. PG helped interpret the results and helped in the writing of the manuscript.

## Notes

### Competing Interest Statement

The authors have declared no competing interest.

### Funding Statement

The author(s) received no specific funding for this work.

